# Interoceptive and metacognitive facets of fatigue in multiple sclerosis

**DOI:** 10.1101/2023.01.23.23284429

**Authors:** Marion Rouault, Inês Pereira, Herman Galioulline, Stephen M. Fleming, Klaas Enno Stephan, Zina-Mary Manjaly

**Affiliations:** Institut du Cerveau et de la Moelle Épinière (ICM), Centre National de la Recherche Scientifique (CNRS), Hôpital Pitié Salpêtrière, 75013 Paris, France; Département d’Études Cognitives, École Normale Supérieure, Université Paris Sciences et Lettres (PSL University), Paris, France; Translational Neuromodeling Unit (TNU), Institute for Biomedical Engineering, University of Zurich & ETH Zurich, Switzerland; Wellcome Centre for Human Neuroimaging, University College London, London, UK; Department of Experimental Psychology, University College London, London, UK; Max Planck UCL Centre for Computational Psychiatry and Ageing Research, University College London, London, UK; Max Planck Institute for Metabolism Research, Cologne, Germany; Department of Neurology, Schulthess Clinic, Zurich, Switzerland; Department of Health Sciences and Technology, ETH Zurich, Switzerland

**Keywords:** confidence, metacognition, fatigue, interoception, perceptual decision-making, computational psychiatry, allostatic self-efficacy, multiple sclerosis.

## Abstract

Numerous disorders are characterised by fatigue as a highly disabling symptom. Fatigue plays a particularly important clinical role in multiple sclerosis (MS) where it exerts a profound impact on quality of life. Recent concepts of fatigue grounded in computational theories of brain-body interactions emphasise the role of interoception and metacognition in the pathogenesis of fatigue. So far, however, for MS, empirical data on interoception and metacognition are scarce.

This study examined interoception and (exteroceptive) metacognition in a sample of 71 persons with a diagnosis of MS. Interoception was assessed by pre-specified subscales of a standard questionnaire (MAIA), while metacognition was investigated with computational models of choice and confidence data from a visual discrimination paradigm. Additionally, autonomic function was examined by several physiological measurements.

Several hypotheses were tested based on a preregistered analysis plan. In brief, we found the predicted association of interoceptive awareness with fatigue (but not with exteroceptive metacognition) and an association of autonomic function with exteroceptive metacognition (but not with fatigue). Furthermore, machine learning (elastic net regression) showed that individual fatigue scores could be predicted out-of-sample from our measurements, with questionnaire-based measures of interoceptive awareness and sleep quality as key predictors.

Our results support theoretical concepts of interoception as an important factor for fatigue and demonstrate the general feasibility of predicting individual levels of fatigue from simple questionnaire-based measures of interoception and sleep.

## Introduction

Fatigue is a central symptom of numerous disorders across medical disciplines (Krupp *et al*., 1989; Wessely, 2001; Chaudhuri & Behan, 2004; Penner & Paul, 2017; Ondobaka *et al*., 2022). Fatigue is fundamentally disabling for patients and profoundly affects their quality of life (Fisk et al., 1994). In general practice, 20% of patients report fatigue as a troubling symptom; this increases significantly in diseases involving dysregulation of the immune system, e.g., autoimmune diseases, cancer, or chronic infections (Dantzer et al., 2014). Finally, fatigue is a common feature of psychiatric disorders. In particular, it constitutes one of the core diagnostic criteria of depression in the Diagnostic and Statistical Manual of Mental Disorders.

In multiple sclerosis (MS), fatigue is a very frequent symptom, with an estimated prevalence of up to 83% (Stuke *et al*., 2009; Kluger *et al*., 2013). Amongst all symptoms in MS, it exerts a particularly profound impact on quality of life (Stuke *et al*., 2009; Penner & Paul, 2017) and represents a critical challenge for clinical management. The pathophysiological mechanisms leading to fatigue in MS are likely diverse (Stuke *et al*., 2009; Penner & Paul, 2017). Unfortunately, so far, we lack any mechanistically interpretable clinical tests that could guide individual treatment. As a consequence, the therapy of fatigue necessarily rests on trial-and-error procedures (Manjaly et al., 2019).

Previous pathophysiological theories of fatigue in MS have focused on a variety of immunological, inflammatory and neurophysiological processes; for a review, see (Manjaly et al., 2019). More recently, a novel perspective on fatigue has been proposed – the “allostatic self-efficacy” (ASE) theory (Stephan *et al*., 2016; Petzschner *et al*., 2017) – that derives from computational theories of brain-body interactions and emphasises the role of two cognitive factors: interoception and metacognition. Interoception – the perception of bodily states – goes beyond the mere registration of bodily sensations, involving an active process of inference based on prior expectations and a model of the body (Seth, 2013; Pezzulo *et al*., 2015; Petzschner *et al*., 2017; Khalsa *et al*., 2018). Metacognition is an umbrella term for “cognition about cognition” (Fleming et al., 2012), comprising evaluation processes by which the brain monitors its own cognitive operations, such as judging the accuracy of perceptual decisions or monitoring the performance of regulatory processes.

The ASE theory builds on a generic mathematical model of brain-body interactions which describes how the brain attempts to control bodily states by monitoring interoceptive surprise (Stephan et al., 2016). Interoceptive surprise is a mathematical quantity that serves as an index of the degree of dyshomeostasis and is computed from prediction errors (PEs), i.e., deviations of actual bodily inputs from the brain’s expectations (under its homeostatic beliefs about the ranges physiological variables should inhabit). Neurophysiologically, interoceptive PEs are reflected by activity in viscerosensory and visceromotor regions (e.g. insula, periaqueductal grey) (Harrison, Köchli, *et al*., 2021).

The ASE perspective proposes that the subjective experience of fatigue arises when, in a situation of persistent dyshomeostasis, the brain arrives at the metacognitive diagnosis that its control over bodily states is failing (Stephan et al., 2016). This metacognitive diagnosis is easily operationalised because the brain only needs to monitor a single quantity – i.e. interoceptive surprise or, equivalently, interoceptive PEs – to detect that dyshomeostasis is not reduced despite regulatory actions (Stephan et al., 2016). Fatigue is thus conceptualised as the experience of low allostatic self-efficacy – an experiential state that reflects the inability to minimise interoceptive PEs by regulatory actions and serves as an imperative signal that suspending any form of action (i.e., rest) is the only remaining option to restore homeostasis.

Empirically, there is initial evidence (direct and indirect) that interoceptive processes are altered in MS (Faivre *et al*., 2012; Rocca *et al*., 2012; Haider *et al*., 2016; Salamone *et al*., 2018; Gonzalez Campo *et al*., 2020) and that changes in interoception are associated with fatigue, in general (Harrison *et al*., 2009) and in MS specifically (Gonzalez Campo et al., 2020). By contrast, the proposed role of metacognition has received little attention so far (but see (Covey *et al*., 2022)). This is due a methodological challenge: although first methods to assess metacognition of interoception are being introduced (Harrison, Garfinkel, *et al*., 2021; Nikolova *et al*., 2022), these are very recent developments (and were not yet available when the present study took place).

Of importance for the present study, recent findings in a large sample from the general population suggest that metacognition of externally directed perceptual processes (exteroception) may also be related to fatigue. According to the ASE theory, this can occur when persistent states of dyshomeostasis (and thus elevated interoceptive PEs) lead to a generalisation of low self-efficacy beliefs beyond interoception; a process putatively associated with the onset of depression (Stephan et al., 2016). Empirically, metacognitive bias (confidence level) during a visual discrimination task was previously found to be related to apathy (Rouault et al., 2018), a construct that overlaps with fatigue. This empirically observed association could reflect domain-independent metacognitive mechanisms, or a possible generalisation of lower confidence to several domains (Seow et al., 2021). This previous finding implies that assessments of exteroceptive metacognition may also become useful in studies of fatigue.

Here, we report results from an observational study that is motivated by the ASE theory (Stephan et al., 2016) and builds on the findings by Rouault et al. (2018). Based on the ASE theory of fatigue, we hypothesised that variability of fatigue levels across persons with MS (PwMS) would be associated with individual differences in interoception, metacognition, and autonomic regulation. To compensate for the current lack of experimental metacognitive probes suitable for directly testing the ASE theory, we resorted to two indirect approaches. First, we used two subscales from an established questionnaire on interoceptive awareness (MAIA) that incorporates aspects of interoceptive awareness, specifically, the tendency not to worry or experience emotional distress with sensations of pain or discomfort, and the experience of one’s body as safe and trustworthy – reflecting the feeling of being in homeostasis and control. As detailed in our pre-specified analysis plan, we expected to find a negative association between these measures and fatigue levels. Second, we investigated whether fatigue might be associated with metacognitive indices that were obtained from for exteroceptive tasks. Given the previously reported association between apathy and metacognitive bias, we hypothesized that metacognitive bias during our exteroceptive tasks would show a negative association with fatigue scores.

Additionally, we conducted several exploratory analyses of links between fatigue and our multimodal data. In particular, using machine learning, we investigated whether individual fatigue scores could be predicted out-of-sample from our measurements and if so, which variables were particularly informative.

## Materials & Methods

### Study participants

We report results from a cross-sectional observational study of adult persons with MS (PwMS) and varying degrees of fatigue. This study was approved by the Ethics Committee of the Canton of Zurich (BASEC number: 2019-00308).

The sample consisted of adult PwMS with an established clinical diagnosis of MS (any clinical subtype). Since our research question concerned the association between fatigue levels and interoceptive/metacognitive factors, we recruited PwMS with different degrees of fatigue, ranging from absent/low to high levels of fatigue. Notably, our observational study did not include a control group of healthy participants without fatigue since this was not necessary for our research question. Moreover, any comparison of groups would have been confounded by the fact that such a control group would have differed from patients in more than one factor, e.g., fatigue, medication, and disease.

To be eligible for the study, PwMS had to fulfil the following inclusion criteria: a diagnosis of MS (according to the revised McDonald criteria (Thompson et al., 2018)) or Clinically Isolated Syndrome, age of ≥18 years, ability to provide written informed consent and adhere to the study protocol. The exclusion criteria were:

- secondary forms of fatigue, e.g., due to anaemia or hypothyroidism,
- use of stimulants (methylphenidate, modafinil) in the last 4 weeks prior to the experiment,
- performing at or below chance level on the session of the metacognition task 1 (see below for task description); chance level is estimated at 55.26% correct (95^th^ percentile of 10,000 simulated responses under a binomial law with *p* = 0.5 and 114 trials),
- reporting the same confidence level on more than 90% of trials of either session of the metacognition tasks (see below for task description), because in that case there is not sufficient variability in confidence ratings for reliably estimating our metacognitive metrics.

Data collection started in June 2019 and was completed in October 2020. 14 participants were included after the World Health Organisation declared the SARS-CoV2 (severe acute respiratory syndrome coronavirus 2) outbreak a global pandemic. In principle, it is possible that pandemic-related stress or biological consequences of SARS-CoV2 infections could act as confounders in that these variables might have influenced both levels of experienced fatigue and interoception and/or metacognition. Since we did not collect any data about pandemic-related stress or infections in our patients, we could not account for these potential confounders in our analyses.

### Sample size

It was difficult to specify a precise power analysis for our study. This was for three main reasons: our observational study tested several hypotheses and with different statistical tests (see below); heterogeneity of pathophysiological mechanisms of fatigue in MS is likely but its degree is unknown (Manjaly et al., 2019); and prior to the beginning of our study, there were no data on the relation of interoception and metacognition to fatigue in MS on which we could have based power calculations. As a general indicator for the required sample size, we therefore assumed a moderate effect size (|r|=0.3) and a nominal significance level of α=0.05, resulting in a target sample of 64 participants to achieve a statistical power of 80% (G*Power Version 3.1) (Faul et al., 2007). For 11 of the participants, we lost parts of the data from the Metacognitive task 1 due to a technical error. We therefore re-measured those participants who were available and recruited a few more participants, resulting in an overall sample size of 71 participants. Note that due to missing data for some of the measurements, each analysis was performed on a subsample of these 71 participants; the exact sample size is reported for each of the analyses and in each of the figures (see Results).

### Study procedures

We tested several hypotheses about statistical relationships between self-report measures of fatigue on the one hand and questionnaire-based measures of interoceptive awareness, measures of autonomic regulation and of metacognition on the other hand while controlling for age, sex and medication as potential confounds.

We provide a detailed account of the hypotheses below. These hypotheses and the statistical procedures to test them were defined prior to data analysis and are described in a prespecified, time-stamped analysis plan available online (https://gitlab.ethz.ch/tnu/analysis-plans/rouault_imefa_analysis_plan). The Results section below indicates whenever procedures deviated from this preregistered analysis plan (e.g., additional analyses).

The study consisted of two sessions on separate days. Session 1 included a clinical interview, a standard neurological and basic neuropsychological examination, and a first computerised behavioural task assessing metacognition. Session 2 comprised the completion of questionnaires (see below), physiological assessments of autonomic function, and a second computerised behavioural task on metacognition. The order of the metacognitive tasks was counterbalanced across participants.

### Measurements

Following an initial clinical examination, the experimental investigation included three types of measurements: questionnaires, measures of autonomic system function, and cognitive tasks (Fig. 1). In addition, several other questionnaire and neuropsychological assessments were conducted for separate research projects. Here, we describe those measures which were of relevance for the research question of the present study.

**Figure 1:**
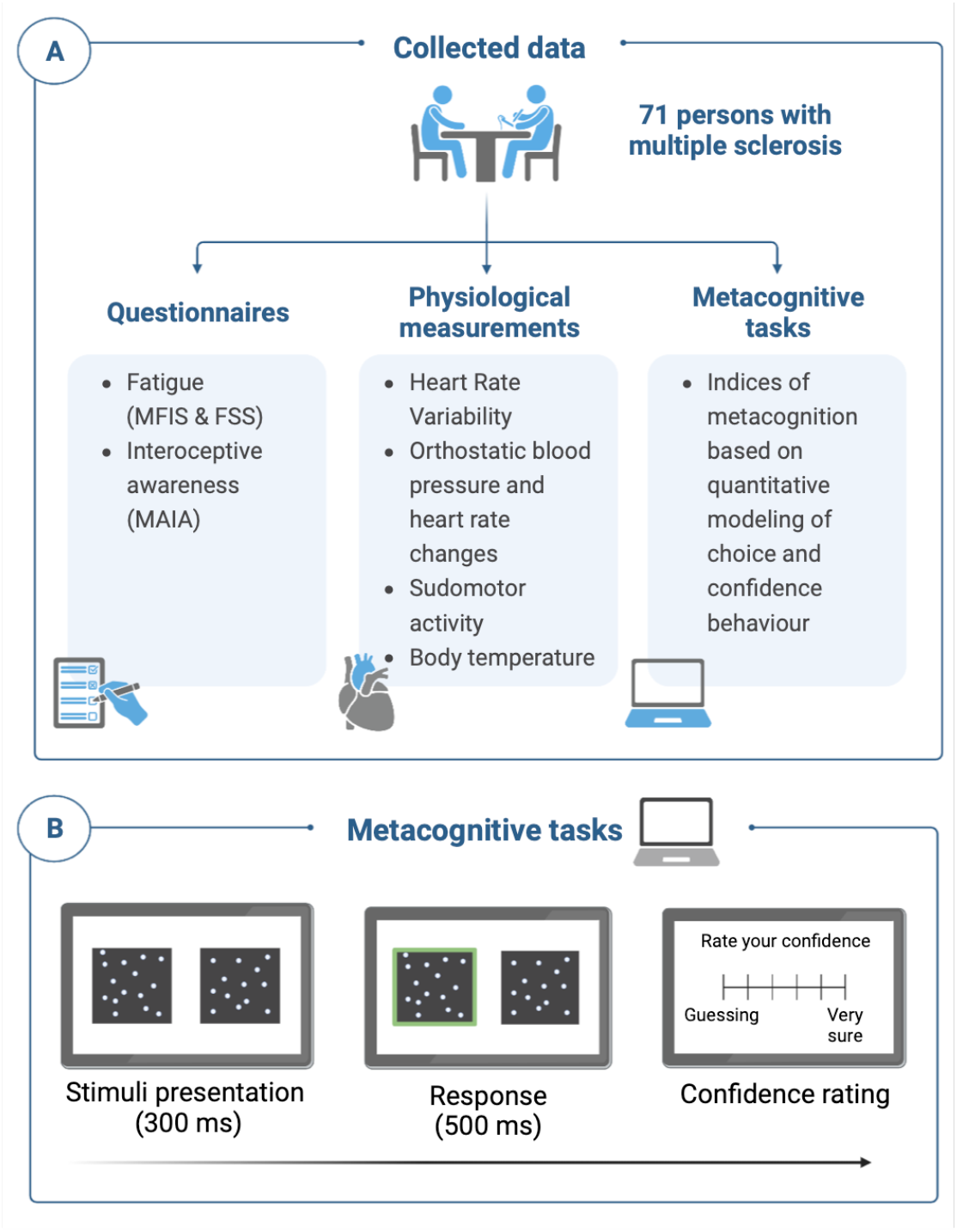
Summary of measurements of metacognition, fatigue and interoception. Participants completed a number of self-report questionnaires, including two fatigue questionnaires, the Modified Fatigue Impact Scale (MFIS) and the Fatigue Severity Scale (FSS). A number of physiological measures of autonomic function were obtained. Green boxes indicate the key variables of interest for metacognition, fatigue and interoception measurements (a full list is provided in the Methods section). Two experimental paradigms assaying metacognition were used to extract summary metrics of decision-making and metacognition. In Metacognition task 1, we used drift-diffusion modeling to characterise participants’ decision-making. We also extracted their confidence level. In Metacognition task 2, we used signal detection theoretic modeling to extract confidence level and metacognitive efficiency which reflects how well a person discriminates between their own correct and incorrect responses (see Methods). Figure created with Biorender.com

#### Clinical examination

Clinical examination included a thorough assessment of neurological status, the Expanded Disability Status Scale, and the Multiple Sclerosis Functional Composite (Rudick et al., 2002).

#### Questionnaires

Our analyses used data from the following questionnaires. To assess individual fatigue levels, we used both the Modified Fatigue Impact Scale (MFIS) (Larson, 2013) and the Fatigue Severity Scale (FSS) (Krupp et al., 1989). This allowed us to verify whether our results were robust to the specific construct of fatigue (sensitivity analysis). To measure the feeling of being in homeostasis and control, we used two subscales (“Not-Worrying” and “Trusting”) from the Multidimensional Assessment of Interoceptive Awareness (MAIA) questionnaire (Mehling et al., 2012). The Hospital Anxiety and Depression Scale (HADS) (Zigmond & Snaith, 1983) was used to assess symptoms of depression. In our context, this is a preferred screening tool for depression since, in contrast to other questionnaires of depressive symptoms, it does not include questions relating to fatigue. The Pittsburgh Sleep Quality Index (PSQI) (Buysse et al., 1989) served to obtain an estimate of sleep. Measures of self-efficacy were obtained by using the MS-Self Efficacy Scale (MSSE) (Chiu & Motl, 2015) and the General Self-Efficacy Scale (GSES) (Schwarzer et al., 1997).

#### Physiological assessments of autonomic function

Concerning physiological tests of autonomic function, the following measurements were obtained: Heart rate variability (HRV, determined by computing the root mean square of successive differences (RMSSD) during deep breathing), changes in blood pressure (ΔBP) and heart rate (ΔHR) when standing up after resting in supine position for 10 minutes, sudomotor activity (Sudoscan, Impeto Medical, France), body temperature (via auricular measurement in the right ear), and sympathetic skin response (SSR). Due to concerns about the quality of SSR measurements, we did not include these data in our analyses.

#### Metacognitive measures

To obtain participant-specific characteristics of metacognition of exteroception, we used two variants of an established experimental paradigm (Rouault et al., 2018). In brief, participants were exposed to a series of visual stimuli – two boxes with different numbers of dots – and, on each trial, were asked to judge which of the two boxes contained more dots. Participants received no feedback but were asked to rate after every decision how confident they were that their decision was correct on a rating scale (Fig. 1). We used a 6-point scale for confidence. Participants performed two variants of the task in two separate sessions described below.

In “Metacognition task 1”, we varied the sensory evidence level (i.e., the dot difference between the two boxes) across trials to cover a large range of decision difficulty levels and model different aspects of the decision formation process using drift-diffusion modelling (DDM). Specifically, two black boxes filled with differing numbers of randomly positioned white dots were presented for 300 ms. One box was always half-filled (313 dots out of 625 positions), while the other box contained an increment of +1 to +70 dots compared to the standard. Participants did 114 trials in 3 blocks of 38 trials. The DDM models a decision via a process of evidence accumulation over time, until a threshold is crossed and the response is elicited. Using the DDM implementation by (Wiecki et al., 2013), we estimated four parameters to characterise this process for each participant: non-decision time (***t***), decision threshold (***α***), baseline drift rate (***v***_**0**_), and the effect of decision evidence on drift rate (***v***_***δ***_). We allowed the evidence level (***δ***) on each decision to affect the drift rate such that:

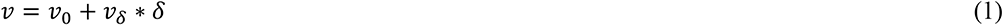

To ensure each participant’s parameter estimates were independent, each participant’s data were fitted separately (Wiecki et al., 2013) using similar procedures as previously reported (Rouault et al., 2018). We also calculated confidence level (also known as “metacognitive bias”) as the average confidence rating provided during the task.

In “Metacognition task 2”, we employed a calibration procedure to maintain a constant level of performance during the experiment and across participants (García-Pérez, 1998). Specifically, we implemented a two-down one-up staircase procedure with equal step-sizes for steps up and down. The staircase was initiated during the practice trials to minimise the burn-in period. Then, participants did 108 trials in 3 blocks of 36 trials. Using hierarchical generative models of recorded responses and confidence ratings based on signal detection theory (Maniscalco & Lau, 2012) (Fleming, 2017), we computed participant-specific indices of metacognitive bias and metacognitive efficiency, two independent metrics of metacognition. Metacognitive bias (or “confidence level”) refers to one’s tendency to rate confidence higher or lower. In contrast, metacognitive efficiency refers to one’s ability to discriminate between correct and incorrect responses, controlling for the influence of task performance (*d’*). This index of metacognitive efficiency is based on meta-*d’* (Maniscalco and Lau 2012), a metric developed in a manner analogous to the classical *d’* index from signal detection theory, but reflecting instead how much information, in signal-to-noise units, is available for metacognitive decisions. Specifically, we extracted for each participant their first-order performance (percent correct and perceptual sensitivity *d’*), metacognitive efficiency (meta-*d’*/*d’*), and metacognitive bias (i.e. confidence level), using the HMeta-d toolbox (https://github.com/metacoglab/Hmeta-d) (Fleming, 2017). This toolbox enables a hierarchical estimation of meta-*d’/d’* over the group (provided in Fig. 3). For regression analyses of the relation between metacognition and other variables (see Analyses B and C below), we employed single-subject Bayesian estimates (Fig. 6 and 7). As a sanity check, we reproduced these regressions using maximum likelihood estimates of meta-*d’*; the results were nearly identical to the Bayesian estimates across all analyses.

### Statistical analysis plan

Our analyses were preregistered in a time-stamped analysis plan available online (https://gitlab.ethz.ch/tnu/analysis-plans/rouault_imefa_analysis_plan). During the analysis, we recognised the need for some adjustments and additional analyses. These deviations from our pre-specified analysis plan are indicated in the Results section below.

We tested statistical relations between self-report measures of fatigue on the one hand and measures of autonomic regulation, metacognition, and questionnaire-based measures of interoceptive awareness on the other hand, while controlling for potential confounds (such as age, sex, medication). We adopted two general analysis approaches:

a. hypothesis-based approaches which tested a priori hypotheses about the relation of fatigue to interoception, autonomic regulation, and metacognition using general linear models (GLM), and
b. exploratory approaches, including principal component analysis (PCA) and elastic net regression.

In our analyses, we used the MFIS questionnaire as a construct of subjectively perceived fatigue. As a sensitivity analysis, we repeated all analyses using the FSS questionnaire as an alternative construct of fatigue.

#### a. Hypothesis-based approach

For testing a set of specific a priori hypotheses, we performed null hypothesis testing using general linear models (GLM), implemented in MATLAB (*glmfit* function). We chose a significance level of α=0.05; where necessary, we corrected for multiple comparisons using a Benjamini-Hochberg (BH) procedure to control the false discovery rate (FDR) at 5%. In all regression analyes, regressors representing potential confounds included age, sex, disease duration (date of data acquisition minus date of initial diagnosis), and medication.

In our pre-specified analysis plan, we had originally envisaged to use dose and drug type as confound regressors. However, the patients in our sample took so many different types of medication that the original plan would have led to a prohibitively large number of regressors in relation to the number of data points. Instead, we thus included two medication-related regressors in our statistical models: one regressor encoding the use of immunomodulatory medication, and a second regressor specifying use of drugs with sedating effects. One additional confound regressor that we had forgotten to specify in our analysis plan was sleep quality (as measured by the Pittsburgh Sleep Quality Index, PSQI). Finally, in one participant, information about the date of initial diagnosis had been entered incorrectly, resulting in a negative disease duration; we replaced this entry with zero.

For all GLMs, we first tested whether they significantly explained variance in general (F-test). If positive, we proceeded to testing several specific hypotheses. We conducted three main analyses and investigated several a priori hypotheses, as described below. All specific hypotheses are formulated as alternative hypotheses (H1) in the context of null hypothesis testing.

### Analysis A: Is fatigue related to measures of interoception and autonomic regulation?

In a first analysis, we modeled the vector of fatigue scores across participants using a GLM with two regressors of interest: the sum of the MAIA subscales 3 (Not-Worrying) and 8 (Trusting), and the first principal component of autonomic measures (HRV, ΔBP and ΔHR (lying vs. standing), sudomotor activity). The reason for using the first principal component (as opposed to including all individual measurements as regressors in our GLM) was that we sought to avoid inflating the number of regressors in relation to our sample size.

We tested the following specific hypotheses:

i. Self-report measures of interoception that relate to the feeling of being in homeostasis and control (i.e. the sum of MAIA subscales 3 and 8) are negatively associated with individual fatigue scores. This hypothesis is a direct prediction from the allostatic self-efficacy model of fatigue and was tested using a one-tailed one-sample *t*-test.
ii. The first principal component of measures of autonomic function is associated with individual fatigue scores. This hypothesis is an indirect prediction from the allostatic self-efficacy model of fatigue (i.e. the model does not directly predict changes in autonomic function but some of the causes for persistent interoceptive surprise the model proposes are connected to autonomic function). In our analysis plan, we had originally specified a negative relationship and thus a directed (one-tailed) t-test, but later realised that a directed relation is difficult to motivate when dealing with principal components of autonomic system measures. We therefore changed this to a more conservative two-tailed one sample t-test.

### Analysis B: Is fatigue related to measures of exteroceptive metacognition?

In this analysis, we examined – separately for the two metacognition tasks – whether fatigue is related to exteroceptive metacognition. For both tasks, the dependent variable was the individual fatigue score (MFIS or FSS, respectively).

For Metacognition task 1, regressors of interest included metacognitive bias (confidence level) as well as estimates of the four DDM parameters characterising the decision formation process during the visual discrimination task: non-decision time, decision threshold, baseline drift rate and the effect of decision evidence on drift rate. We tested the following hypothesis using a one-tailed one-sample *t*-test:

(iii) Metacognitive bias (confidence level) is negatively associated with fatigue. This hypothesis is based on the previous result by Rouault et al. (2018) who found that apathy, a construct related to fatigue, was negatively associated with confidence level in the same visual discrimination task as used in this study.

Metacognition task 1 not only allowed us to examine potential shifts in metacognition, but also shifts in other cognitive performance features (here, the decision formation process). We also examined the associations between DDM parameters and fatigue but had no specific directional hypotheses (Rouault et al., 2018; Ulrichsen et al., 2020).

For Metacognition task 2, regressors of interest included metacognitive bias (confidence level) and metacognitive efficiency (meta-*d’*/*d’*). For metacognitive bias, we also added accuracy as a regressor of no interest. Although accuracy is matched across participants by design, integrating it in the model allow us to fully isolate effects of bias from any remaining effects of performance fluctuations around the target value from the staircase. We tested the following hypotheses using one-sample t-tests:

(iv) metacognitive efficiency (meta-*d’*/*d’*) is associated (unspecified direction) with fatigue.
(v) metacognitive bias (confidence level) is negatively associated with fatigue. This is the same as hypothesis (iii), and we expected to find consistent relationships across both metacognition tasks.

### Analysis C. Are measures of interoception and autonomic regulation related to measures of metacognition?

We used two different GLMs with the same design matrix as in analysis B but concerning different dependent variables. The first GLM attempted to explain variance in MAIA scores by metacognitive indices (confidence level, metacognitive efficiency), whereas in the second GLM, the dependent variable was the first principal component obtained from the physiological measures of autonomic regulation (HRV, ΔBP and ΔHR lying vs. standing, sudomotor activity). We tested the following hypotheses:

(vi) metacognitive indices are associated with individual levels of questionnaire-based interoceptive awareness that relate to the feeling of being in homeostasis and control (sum of MAIA subscales 3 [Not-Worrying] and 8 [Trusting]);
(vii) metacognitive indices are associated with the first principal component of measures of autonomic function.

#### b. Exploratory analyses

First, we examined an association between ‘global’ confidence (i.e. self-efficacy) and ‘local’ confidence, i.e. the task-specific confidence level within the context of the visual discrimination paradigms used here. This exploration is relevant because metacognition can operate across different levels of abstraction, from ‘local’ confidence in individual perceptual decisions, to global beliefs about general abilities such as self-efficacy (Rouault et al., 2019). Given known associations between feelings of self-efficacy and feelings of confidence, we initially examined whether confidence level from the Metacognitive Task 2 correlated with either of the two self-efficacy questionnaires in our study (MS Self-Efficacy Scale and General Self-Efficacy scale).

Second, we performed a multivariate linear regression analysis with nested cross-validation and elastic net regularisation (Zou & Hastie, 2005). This analysis included all autonomic, questionnaire-based interoceptive and task-based metacognitive measurements and aimed at identifying the most meaningful predictor(s) of fatigue scores without a priori hypotheses or preselection of regressors. Elastic net regularisation introduces two penalty terms to the ordinary least squares objective function that combine properties of lasso (L1 penalty) and ridge regression (L2 penalty), allowing selection of a solution with particular properties. In particular, for problems with few data points compared to the number of regressors, this avoids overfitting, thus enhancing the accuracy of the predictors, through automatic variable selection and shrinkage of large regression coefficients.

In our application of elastic net regression implemented in *scikit-learn*, we z-scored the predictors (regressors) and mean-centred the outcome variable (fatigue scores). As in Rouault et al. (2018), we implemented ten-fold cross-validation (CV), with nested cross-validation for tuning the hyperparameters. The data were randomly split into 10 sets (folds). A model was then generated based on 9 training folds, and applied to the remaining independent validation set. Each fold served as the validation set once, resulting in 10 different models and predictions. Nested cross-validation involved subdividing the 9 training sets (i.e., 90% of the sample) into a further 10 folds (“inner” folds). Within these 10 inner folds, 9 were utilized for training a model over a range of 10 alpha (0.01–1) and 10 lambda (0.01–100) values, where alpha is the complexity parameter and lambda is the regularisation coefficient. This resulted in a model fit for the inner test set for each possible combination of alpha and lambda. The best fit over all 10 inner folds for each combination of alpha and lambda was then used to determine the optimal parameters for each outer fold. We tested the significance of regression coefficients using permutation tests with 1,000 permutations.

### Data and code availability

All participants were asked for consent that their pseudonymised data could be shared; for those participants who agreed (68 out of 71 participants), the data are available on the ETH Research Collection (www.research-collection.ethz.ch/handle/20.500.11850/609819). The MATLAB code for the GLM analyses is available at https://www.github.com/marionrouault/imefa/ and the Python code for the elastic net analysis is available at https://gitlab.ethz.ch/hermang/rouaultetal_fatigue_elastic_net.

## Results

### Characteristics of participants

Overall, 71 participants were included in the study. 68 participants had a relapsing-remitting form of MS, one participant had a primary progressive MS, and two participants had a secondary progressive MS. We first examined age, sex, medication status and disease duration in our sample (Fig. 2). Mean age was 42.4 years old (median=43, min=19, max=67) and the gender distribution was strongly skewed (61 female, 9 male, 1 missing data). Most participants (62 persons) were on medication (17 fingolimod, 14 dimethyl fumarate, 8 ocrelizumab, 7 natalizumab, 5 interferon beta-1a, 3 glatiramer acetate, 2 alemtuzumab, 2 teriflunomide, 1 rituximab, 1 peginterferon beta-1a). Eleven participants took supplementary medication (e.g., benzodiazepines) with sedating side effects. The mean disease duration was 7.6 years (median=6.2, min=0, max=31.2) (Fig. 2A).

**Figure 2:**
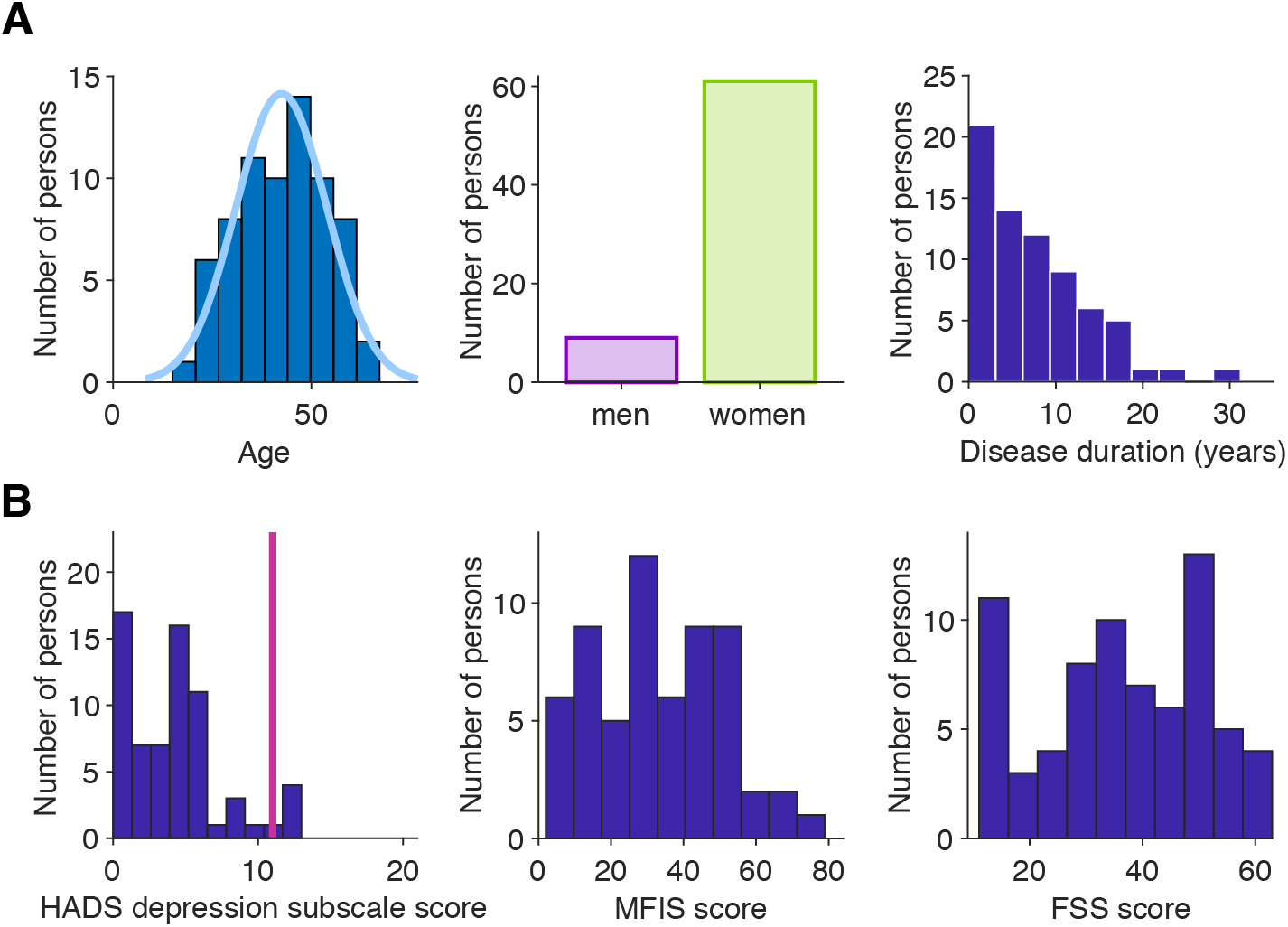
Characteristics of the sample of persons with MS. **A)** Demographic metrics of participants: age, sex, and disease duration histograms. **B)** Left panel: distribution of the Depression subscale of the Hospital Anxiety and Depression Scale (HADS) scores in our sample (N=68 available measures). HADS depression subscale score varies from 0 to 21. The vertical line indicates the standard cut-off considered as clinically relevant; this applied to 5 participants. Middle and right panel: fatigue scores as measured by the Modified Fatigue Impact Scale (MFIS) (N=61 available measures) and by the Fatigue Severity Scale (FSS) (N=71 available measures).

Our inclusion criteria did not impose any constraints with regard to levels of fatigue and depressive symptoms. Concerning fatigue levels in our sample, according to the Modified Fatigue Impact Scale (MFIS), participants exhibited significant levels of fatigue with an average score of 33.1 (SD=18.3). Likewise, participants had an average score on the Fatigue Severity Scale (FSS) of 36.6 (SD=14.8) (Fig. 2B). Despite the visually different shapes of their distributions, scores on these two fatigue scales were strongly correlated, as expected (Spearman ρ=0.87, *p*=5.6×10^-20^).

According to the Hospital Anxiety and Depression Scale (HADS; N=68 available measures), study participants showed a fairly moderate degree of depressive symptoms (average score on the depression subscale of the HADS: 4.2, SD=3.3) (Fig. 2B). Five participants reached a score indicative of probable depression according to established thresholds (Zigmond & Snaith, 1983).

### Estimating indices of (exteroceptive) metacognition

As a probe of exteroceptive metacognition, we used a well-validated metacognition task in the domain of visual discrimination (Rouault et al., 2018), in order to extract key metacognitive metrics using drift-diffusion and signal detection theoretic models (see Methods). In Metacognition task 1, participants (N=68 available measures) achieved a mean performance above chance level of 69% correct (min=46%, max=86%) (Fig. 3A). As expected, increased perceptual difficulty (i.e., smaller difference in the number of dots between left and right boxes) was associated with lower performance and longer response times (RTs) (difference between first and last difficulty bin: *t_67_*=-15.4, *p*=1.2×10^-23^ (accuracy), *t_67_*=4.8, *p*=1.0×10^-5^ (RTs)) (Fig. 3B). Participants stated higher confidence for correct than incorrect decisions (*t_67_*=9.3, *p*=1.056×10^-13^), indicating a significant degree of metacognitive sensitivity (Fig. 3A). In Metacognition task 2 (N=67 available measures), participants achieved a mean performance above chance level of 75% correct (min=68%, max=81%), close to the level of 71% correct targeted by the staircase procedure (Fig. 3C). Again, participants indicated higher confidence for correct than incorrect decisions (*t_66_*=9.5, *p*=4.78×10^-14^) (Fig. 3D), and we found a group-level metacognitive efficiency of 0.79 (mean H-Mratio from hierarchical fit with satisfactory convergence of ^R^^ =1.0005).

**Figure 3:**
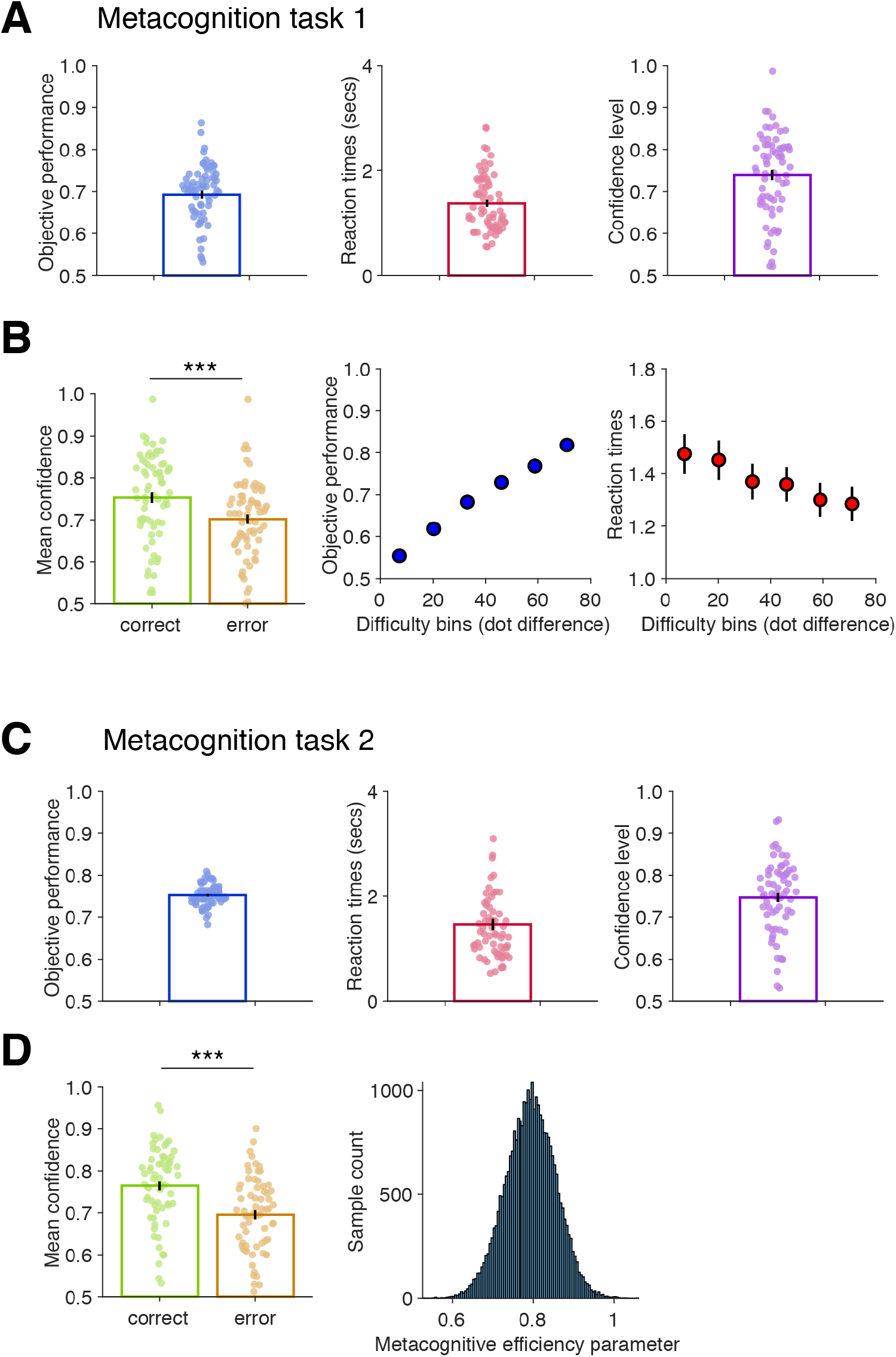
Participants’ behaviour on the metacognitive tasks. **A**) Participants’ performance, response times, and confidence level in Metacognition task 1. Bars and error bars indicate mean and standard error of the mean (SEM); dots indicate individual data points (N=68 available measures). **B**) Left panel: Participants’ confidence for correct and incorrect decisions. Right panel: Mean perceptual performance (left panel) and mean response time (right panel) in each of the six difficulty bins, determined by the difference in number of dots between left and right boxes. Error bars indicate SEM (N=68). **C**) Participants’ performance, response times, and confidence level in Metacognition task 2. Bars and error bars indicate mean and SEM and dots indicate individual data points (N=67 available measures). **D**) Left panel: Participants’ confidence for correct and incorrect decisions. Right panel: Group-level metacognitive efficiency (H-Mratio) distribution estimated hierarchically (see Methods).

### Dimensionality reduction of physiological measures

In order to keep our regression models as parsimonious as possible, we aimed to obtain a low-dimensional summary of the physiological measures. To this end, we used principal components analysis (PCA) and computed the first principal component of physiological measurements, as pre-specified in our analysis plan (see Methods). We also examined the pairwise correlations of our measurements to better understand the covariance structure (Fig. 4A). As expected, sudomotor activity was correlated between hands and feet (ρ=0.62, *p*=4.38×10^-8^), and ΔBP was correlated between systolic and diastolic measurements (ρ=0.35, *p*=0.0048). The other physiological measurements of interest (HRV, ΔHR, and sudomotor activity) were reasonably uncorrelated (Fig. 4A). Applying PCA to all physiological measurements (Fig. 4B), we found that the first principal component (PC1) explained 42.2% of the variance in measurements of autonomic function.

**Figure 4:**
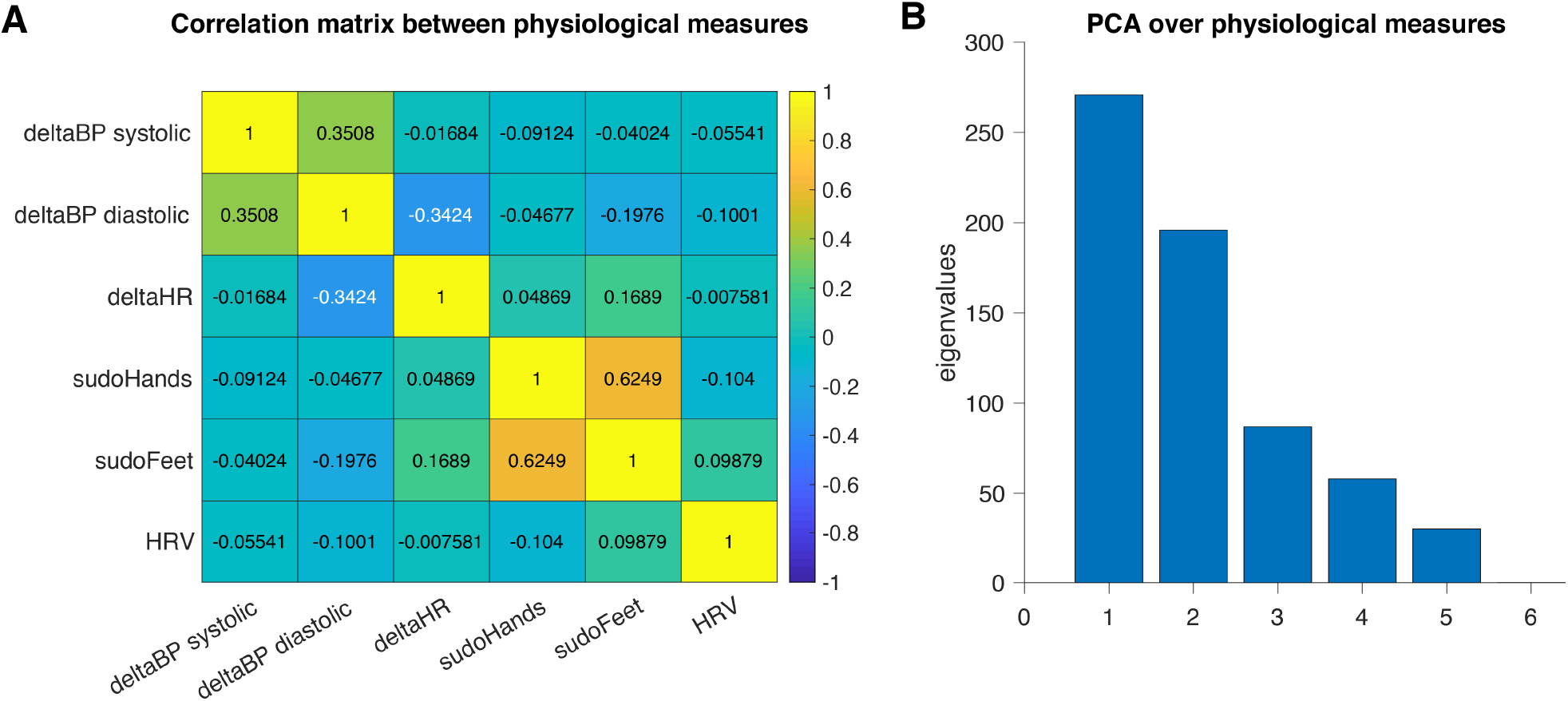
Physiological measures of homeostatic regulation. **A**) Correlation matrix of participants’ heart rate variability (HRV, computed as RMSSD during deep breathing), systolic and diastolic ΔBP and ΔHR (standing up after resting in supine position for 10 minutes), and sudomotor activity (averaged over hands and feet). **B**) Eigenvalues obtained for each of the inputs from a principal component analysis (PCA).

With all of our metrics established, we next turned to the hypotheses pre-specified in our analysis plan.

### Analysis A: Is fatigue related to measures of interoception and autonomic regulation?

Starting with hypothesis A (see Methods), we examined whether fatigue scores were associated with self-report, questionnaire-based measures of interoceptive awareness that relate to the feeling of being in homeostasis and control and with the first principal component (PC1) of physiological measurements of autonomic function. Controlling for a number of potential confounds (see Methods), overall, the regression model explained a significant amount of variance in fatigue (MFIS) scores (F-test: *p*=0.0372; N=53 available measures). We found a significant negative relation of self-report measures of interoception (MAIA subscales 3 and 8) with fatigue scores (*t_52_*=-2.79, one-tailed *p*=0.0078 uncorrected, *p*=0.025 FDR-corrected), but failed to find a significant association with PC1 (*t_52_*=0.41*, p=*0.68) (Fig. 5). The pattern of findings was similar when using the FSS questionnaire for fatigue instead of MFIS. Again, the regression model overall explained a significant amount of variance in FSS scores (F-test: *p*=7.9×10^-4^; N=63 available measures). Furthermore, there was again a significant negative association of fatigue levels with MAIA scores (*t_62_*=-3.04, one-tailed *p*=0.0036 uncorrected, *p*=0.05 FDR-corrected), but not with PC1 (*t_62_*=-0.03, *p*=0.97) with fatigue (FSS) scores.

**Figure 5:**
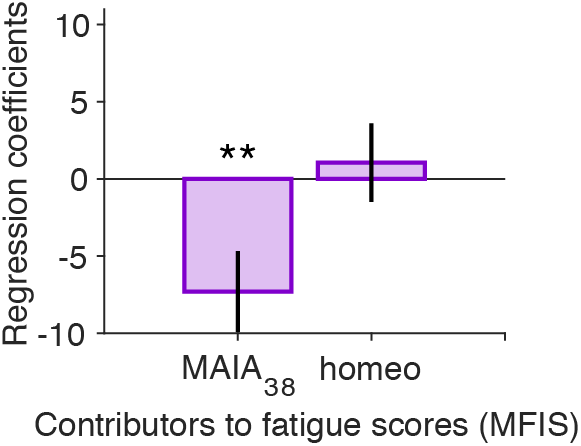
Association of fatigue levels to measures of interoception and autonomic regulation (Analysis A). Regression analysis of MFIS values, with MAIA subscales relating to the feeling of being in homeostasis and control (‘MAIA_38_’) and autonomic function measures reflecting the integrity of homeostatic regulation (‘homeo’) (see Methods). Regressors of no interest include age, sex, immunomodulatory medication, medication with sedative effects, disease duration, and sleep quality as measured by the PSQI questionnaire. Error bars are the standard errors of regression coefficient estimates (N=53 available measures). **p<0.01 uncorrected, p<0.05 FDR-corrected; p-values from one sample t-tests against zero on regression coefficient. Note that fatigue scores based on the FSS questionnaire provided very similar results (see main text).

Given the surprising absence of an association between fatigue and physiological measurements of autonomic function, we conducted several control analyses, examining in particular whether using the first principal component as a summary of the various autonomic measurements may have been an inadequate choice. These control analyses, which are reported in the Supporting Information, confirmed that in our particular sample, a significant association between autonomic function measures and fatigue is not found.

Altogether, these findings indicate that self-report (questionnaire-based) measures of interoception that reflect the feeling of being in homeostasis and control were significantly related to fatigue, whereas this was not the case for physiological measures of autonomic function.

### Analysis B: Is fatigue related to measures of exteroceptive metacognition?

The second part of our pre-specified analyses focused on the relationship between fatigue and (exteroceptive) metacognition. Here, the most important question – which we tested twice, using data from both metacognition tasks – was whether fatigue would show a negative association with metacognitive bias (confidence level). This hypothesis was based on previous findings by Rouault et al. (2018) who found this association for a fatigue-related construct (i.e., apathy) in the Metacognition task 2.

First, using Metacognition task 1 and parameter estimates from a drift-diffusion model of the decision-making process (see Methods), we found that our regression model did not significantly explain variance in fatigue (MFIS) scores (F-test: *p*=0.1380, N=52 available measures). Contrary to our expectation, we failed to find a significant association between metacognitive bias and fatigue (one-tailed t-test, *t_54_*=0.03, *p*=0.97). Amongst the hierarchical drift diffusion model (HDDM) parameters, we found that neither the baseline drift rate (*v*_0_) (*t_54_*=-1.90, *p*=0.0646) nor the effect of decision evidence on drift rate (*v*_δ_) (*t_54_*=- 1.42, *p*=0.163) were significantly associated with fatigue. For the other two parameters, decision threshold and non-decision time, we again failed to find a significant relation with fatigue (both *t_54_*>-1.17, both *p*>0.25) (Fig. 6A).

**Figure 6:**
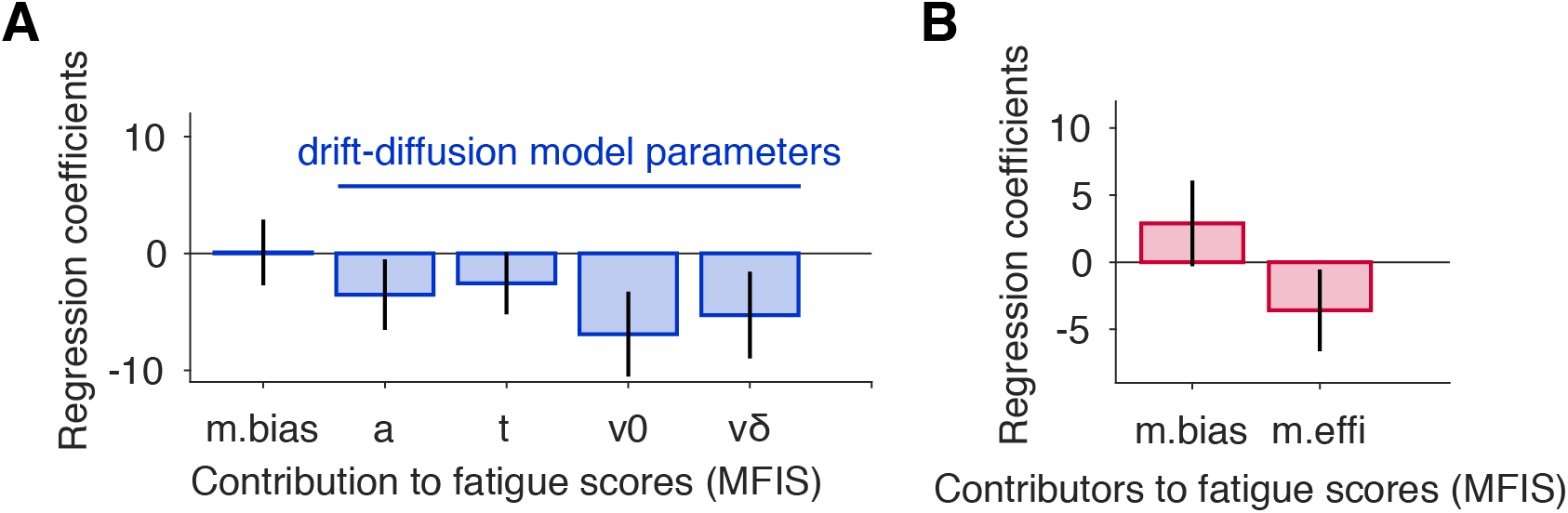
Association of fatigue levels to measures of metacognition (Analysis B). Analyses of the relationship between metacognition and fatigue scores (MFIS questionnaire). **A**) From Metacognition task 1, we included as regressors metacognitive bias (‘m.bias’) alongside four drift-diffusion model parameter estimates characterising the perceptual decision-making process: decision threshold (‘a’), non-decision time (‘t’), drift rate (*v*_0_ and *v_δ_*) (see Methods) (N=52 available measures). **B**) From Metacognition task 2, we included as regressors metacognitive bias (‘m.bias’) and metacognitive efficiency (meta-d’/d’) (‘m.effi’). (N=56 available measures). In all models, regressors of no interest included age, sex, immunomodulatory medication, medication with sedative effects, disease duration, and sleep quality (PSQI questionnaire). Error bars are the standard errors of regression coefficient estimates.

Going beyond our pre-specified analyses, we reasoned that unpacking the decision process into the four HDDM parameter estimates might have led to an overparameterised regression model, and that replacing them by general decision accuracy (which the drift diffusion model seeks to characterise) could result in a more parsimonious model. Therefore, we ran an alternative model replacing the four HDDM parameters with accuracy instead (F-test: *p*=0.0271). We found that lower accuracy was related to fatigue (*t_54_*=-2.45, *p*=0.0176 uncorrected, *p*=0.025 FDR-corrected), again in the absence of a significant association between metacognitive bias (confidence level) and fatigue (one-tailed t-test, *t_54_*=0.679, *p*=0.50).

Second, using Metacognition task 2 and a Bayesian model based on signal detection theory (Fleming, 2017), we extracted metacognitive bias (confidence level) and metacognitive efficiency (meta-*d’*/*d’*) and examined their link with fatigue using multivariate regression (see Methods). The regression model overall did not explain a significant amount of variance (F-test, *p*=0.2122), we found that neither metacognitive bias (one-tailed t-test, *t_55_*=0.91, *p*=0.185) nor metacognitive efficiency (two-tailed t-test, *t_55_*=-1.18, *p*=0.24) were significantly associated with fatigue (Fig. 6B). Deviating from our specified analysis plan, we did not include accuracy in the model because it is already partly taken into account in the calculation of metacognitive efficiency (via *d’*). However, we also implemented the same model by adding the average evidence level for each individual from the staircase procedure, a proxy for individual perceptual difficulty, which provided consistent results. This model was significant (F-test: *p*=0.0417), with neither metacognitive bias (one-tailed t-test, *t_55_*=0.75, *p*=0.23) nor metacognitive efficiency (two-tailed t-test, *t_55_*=-1.22, *p*=0.23) being significantly associated with fatigue, in contrast to significant effects of average evidence level (*t_55_*=2.60, *p*=0.0123) on fatigue.

### Analysis C. Are measures of interoception and autonomic regulation related to measures of metacognition?

Finally, we examined our third pre-specified set of hypotheses regarding associations of interoception and autonomic measures, respectively, with metacognition (Analysis C). First, using multivariate regression, we examined whether self-report, questionnaire-based measures of interoceptive awareness (sum of MAIA subscales 3 and 8) were related to metacognitive indices (see Methods). We found that the model did not significantly explain more variance than a null model (F-test: *p*=0.60), and that none of the metacognitive regressors significantly explained MAIA subscale scores (all abs(*t_65_*)<0.69, all *p*>0.49; N=66 available measures; Fig. 7A).

**Figure 7:**
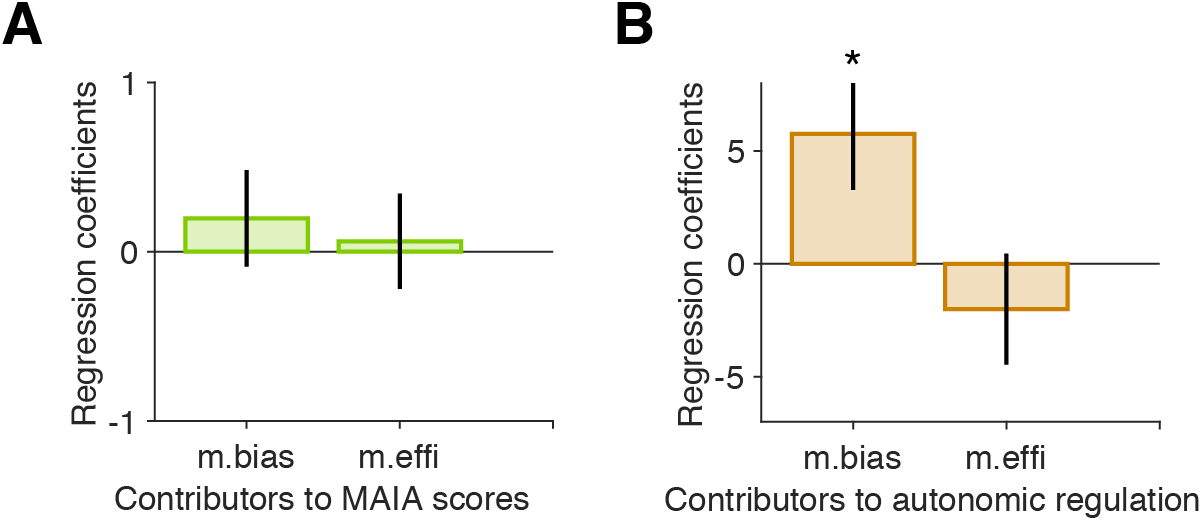
**Association between measures of interoception and autonomic regulation and measures of metacognition (Analysis C). A**) Regression analysis of the contribution of metacognitive bias and metacognitive efficiency to explaining a self-report measure of interoception (MAIA subscales, see Methods) (N=66 available measures) **B**) Regression analysis of the contribution of metacognitive bias and metacognitive efficiency to explaining a physiological measure of autonomic function (PC1, see Methods). (N=61 available measures). In both models, regressors of no interest include age, sex, immunomodulatory medication, medication with sedative effects., disease duration, and sleep quality as measured by the PSQI questionnaire. Error bars are the standard errors of regression coefficient estimates. *p<0.05, uncorrected p-values from one sample t-test against zero on regression coefficient. Error bars are the standard errors of regression coefficient estimates.

Second, we examined, using multivariate regression with the first principal component of physiological measures of autonomic function (PC1) as dependent variable, whether PC1 was related to metacognitive indices. Although the overall model did not explain significantly more variance than a null model (F-test: *p*=0.1213), we did find a significant association between metacognitive bias (confidence level) and PC1 (*t_60_*=2.32, *p*=0.0243) (Fig. 7B).

Altogether, these results suggest that exteroceptive metacognitive bias is associated with physiological measures of autonomic function, but not with aspects of interoceptive awareness related to the feeling of being in homeostasis and control (summed MAIA subscales 3 and 8).

### Elastic net regression: predicting fatigue from all available measurements

The analyses described above used classical within-sample multiple regression with carefully pre-selected regressors, including the use of dimensionality reduction (PCA), in order to examine relationships between fatigue, metacognition, and interoceptive markers in a hypothesis-driven manner. In a subsequent and more exploratory step (but part of our pre-specified analysis plan), we performed a regression analysis where we sought to predict fatigue from all of the available interoceptive, physiological, and metacognitive measurements as well as additional variables (e.g., sleep; see Methods). This analysis was only possible for those participants where measures of all 15 variables were available (N=52 participants for MFIS, N=62 for FSS).

In order to avoid overfitting and obtain out-of-sample predictions, we used elastic net regularisation (Zou & Hastie, 2005) together with ten-fold nested cross-validation. We used permutation tests to examine whether model predictions as well as the contribution of specific regressors were significantly above chance. Specifically, we derived null distributions based on the mean squared error (MSE) for model predictions and based on the regression coefficients for individual regressors. In either case, 1,000 permutations were used to create the null distribution.

The regression model was able to predict individual MFIS scores well above chance (*p*=0.003; Fig. S2). Out of the 15 regressors, two showed large regression weights, and both were significant predictors of MFIS scores: self-report measures of interoceptive awareness, i.e., summed scores of MAIA subscales 3 and 8 (regression weight = -5.49, *p*=0.002), and sleep quality (regression weight = 4.83, *p*=0.001). While these results confirm the relation between fatigue and interoceptive measures, it is noteworthy that they were now obtained in the presence of all other variables. Moreover, the use of cross-validation moves the analysis from *explaining* fatigue scores (i.e., within-sample associations) towards *predicting* them out-of-sample. Specifically, the model in the current analysis can predict MFIS scores of “unseen” individuals with a median absolute error of 13.59 (for comparison, MFIS scores are on a scale from 0 to 84).

Turning to FSS as an alternative fatigue score, again the model’s predictions were significantly above chance (*p*=0.003; Fig. S3). The same regressors as for MFIS were significant predictors of fatigue: self-report measures of interoceptive awareness (MAIA subscales 3 and 8) (regression weight = -3.47, *p*=0.001) and sleep quality (regression weight = 5.05, *p*=0.001). The overall model could predict individual FSS scores out-of-sample with a median absolute error of 10.19 (for comparison, FSS scores are on a scale from 9 to 63).

Finally, it is worth mentioning that, for MFIS scores, two of the physiological variables (ΔHR and HRV) were also significant predictors (ΔHR: *p*=0.019; HRV: *p*=0.031). However, this finding did not generalise across questionnaires: for FSS scores, neither variable was a significant predictor (ΔHR: *p*=0.763; HRV: *p*=0.749).

In conclusion, as for the separate regression analyses above, the choice of fatigue questionnaire did not impact our results: both MFIS and FSS fatigue scores could be predicted with highly significant accuracy, and the same variables (self-report on interoception and sleep quality) were important for this prediction.

### Relationship between local and global confidence

Finally, as pre-specified in our analysis plan, we examined possible associations between ‘global’ confidence (here measured as self-efficacy) and ‘local’ confidence, the task-based confidence level, by examining the correlation between metacognitive bias (confidence level) from Metacognition task 2 and either of the two self-efficacy questionnaires that participants had completed (General Self-Efficacy Scale, GSES, and MS Self-Efficacy Scale, MSES). For GSES, the Pearson correlation coefficient was *ρ*=0.11 (*p*=0.37) and for MSES *ρ*=0.067 (*p*=0.59) (N=66 available measures in both cases). The correlations are visualised in Fig. 8. These findings suggest that, in our particular sample and for the task we used, task-based metacognitive confidence and general self-efficacy beliefs were not significantly related.

**Figure 8:**
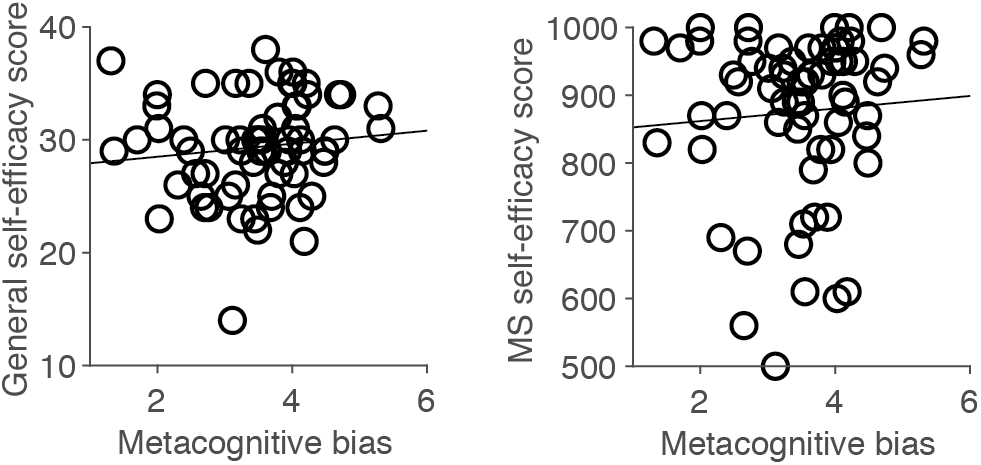
Associations between ‘local’ and ‘global’ aspects of metacognition. Correlations between task-based measure of metacognitive bias (‘local’ confidence level) and global measures of confidence as indexed by the general self-efficacy scale (GSES, left panel) and the MS self-efficacy scale (right panel).

## Discussion

This study examined questionnaire-based interoceptive awareness, autonomic function, and (exteroceptive) task-based metacognition in a sample of 71 PwMS. We tested a number of concrete hypotheses jointly inspired by the theory of allostatic self-efficacy (ASE) (Stephan et al., 2016) and by recent findings on links between exteroceptive metacognition and apathy in a general population sample (Rouault et al., 2018). In brief, our study found the expected association of interoceptive awareness with fatigue (but not with exteroceptive metacognition) and an association of autonomic function with exteroceptive metacognition (but not with fatigue). Furthermore, a machine learning analysis (based on elastic net regression) showed that individual fatigue scores could be predicted out-of-sample from our measurements, with questionnaire-based measures of interoceptive awareness and sleep quality having a particular relevance for prediction.

To the best of our knowledge, our study is novel in at least three ways. It is the first to explore the utility of (exteroceptive) measures of metacognition for investigating fatigue. Second, it achieves successful out-of-sample prediction of individual fatigue scores in MS from simple questionnaire-based measurements. Third, when examining links between questionnaire-based interoception and fatigue, it is the first study that assesses interoceptive awareness in MS using a validated questionnaire (MAIA) focusing on those aspects of interoceptive awareness that relate to the feeling of being in control and homeostasis.

How do our results relate to previous findings in the literature? Several previous studies have provided indirect evidence for an association between altered interoception and fatigue in MS. This rested on showing that PwMS exhibit structural/functional changes in interoceptive brain regions, such as the insula and anterior cingulate cortex (ACC) (e.g., (Faivre *et al*., 2012; Rocca *et al*., 2012; Haider *et al*., 2016; Salamone *et al*., 2018)), and demonstrating associations of such changes with fatigue (Andreasen et al., 2010; Pardini et al., 2015). This form of evidence for an association between altered interoception and fatigue is highly valuable but only of an indirect nature because areas like the ACC and the insula are also involved in other cognitive functions. Direct assessments of interoception in MS are rare so far. The only exception we are aware of is a recent study which showed that, in comparison to healthy controls, PwMS with fatigue (but not PwMS without fatigue) exhibited significant differences on a heartbeat detection task (in addition to changes in grey matter volume and functional connectivity of the insula) (Gonzalez Campo et al., 2020). Our study used specific subscales of the MAIA questionnaire (pre-specified in our analysis plan) to assess aspects of interoceptive awareness that are of particular relevance for the ASE theory. We are not aware of any previous study that used the MAIA or another validated interoception questionnaire in MS.

Concerning the relation between autonomic nervous system function and fatigue in MS, most previous studies have investigated cardiac measures of autonomic dysfunction, e.g., with regard to heart rate variability (Flachenecker et al., 2003). As summarised in a recent systematic review of cardiac autonomic function in MS, most of these studies have reported a relation between cardiac autonomic dysfunction and fatigue, although the degree and the nature (i.e. which measurement) of this relation varied considerably across studies (Findling *et al*., 2020). In our analyses, we did not find any association between measures of autonomic function and fatigue when applying dimensionality reduction (PCA) prior to regression analysis. When using all physiological measures as regressors in a regularised regression model (elastic net), we found that two physiological variables (ΔHR and HRV) did predict MFIS scores; however, this finding could not be replicated for FSS scores and should therefore be treated with caution. We conducted a number of control analyses (see Supporting Information) which, however, failed to reveal an obvious reason for the absence of the expected association. One remaining possibility is that although our sample size was relatively large (N=71 participants), this may still not have been large enough to detect associations of small effect size.

As specified in our analysis plan, we also conducted an exploratory analysis, applying a machine learning approach (elastic net regression with ten-fold nested CV) to all our measurements. It showed that fatigue scores of individual participants can be predicted from questionnaire, physiological and behavioural measurements, with a median absolute error of 13.59 for MFIS and 10.19 for FSS. Furthermore, questionnaire-based measures of interoceptive awareness and sleep quality played a particularly important role for this prediction. This finding is important in two ways. First, this machine learning approach goes beyond classical within-sample statistical analyses: by combining regularisation with cross-validation, it doubly protects against overfitting and enables us to include all measurements within a single regression model. This allows our analysis to account for several potential confounders, thus decreasing the probability that the observed predictive relationship may have been driven by a third variable. The finding that sleep quality by itself is negatively related to fatigue is not surprising and has been demonstrated before (e.g. (Nociti *et al*., 2017)); however, our analysis provides new evidence for the strength of this relationship by examining it in the simultaneous presence of many other explanatory variables and in an out-of-sample prediction context. Second, patients are often frustrated by the lack of objective tests that provide objective confirmation of their subjective experience of fatigue. An important clinical goal is therefore to predict the presence and degree of fatigue from other measurements. Clearly, our current study does not present a solution to this long-standing problem because the predictive variables it identifies (questionnaire-based interoceptive awareness and sleep quality) are based on self-reports themselves and because the sample size is too small to for establishing a precise prediction tool. Nevertheless, to our knowledge, it represents the first demonstration that predicting individual fatigue levels out-of-sample is possible at all for MS.

While numerous studies exist which, in a variety of contexts and disorders, used machine learning to predict individual fatigue levels from behavioural or physiological data (Baykaner *et al*., 2015; Mun & Geng, 2019; Luo *et al*., 2020; Bafna *et al*., 2021; Jiang *et al*., 2021; Pinto-Bernal *et al*., 2021; Yao *et al*., 2021; Zeng *et al*., 2021), only two of these studies have concerned MS (Ibrahim *et al*., 2020; 2022). Additionally, like the vast majority of studies, these two MS-specific studies did not actually predict *fatigue* (the subjective experience) but predict *fatiguability* (the observable decrease in performance during physiologically or cognitively demanding tasks) (Kluger et al., 2013) (sometimes, fatigue and fatiguability are also referred to as “trait fatigue” and “state fatigue”, respectively (Cehelyk *et al*., 2019)). Generally, to our knowledge, only two previous studies – concerning fatigue during cancer treatment and in HIV, respectively; (Zuñiga *et al*., 2020; Kober *et al*., 2021) – have attempted to predict fatigue, all other studies have reported predictions of fatiguability. While fatiguability is also a clinically very important topic, it is important to distinguish these two concepts, not least because different types of pathophysiological explanations exist for fatigue and fatiguability (Manjaly et al., 2019).

Our study has a number of notable strengths and limitations. Beginning with its strengths, our results derive from a preregistered analysis plan in which all hypotheses and analysis procedures were specified before the data were touched. Any deviations from this analysis plan were described in the Methods and Results sections above. Second, we examine (exteroceptive) metacognition using an established behavioural paradigm and a computational (hierarchical Bayesian) model (Fleming, 2017; Harrison, Garfinkel, *et al*., 2021). Third, a sensitivity analysis – i.e., comparing our results across two separate fatigue questionnaires (MFIS and FSS) – demonstrated that our findings did not depend on a specific construct of fatigue. It could be interesting in future work to examine relationships between different dimensions of fatigue (e.g. physical vs. cognitive) and measures of interoception/metacognition. However, in this study, we did not conduct separate analyses for different dimensions of fatigue because the ASE theory (which guided our analyses) does not make any predictions in this regard so far. Finally, as mentioned above, using machine learning, we could take into account many potential confounders, despite a limited sample size, and demonstrate that individual fatigue levels can be predicted, out-of-sample and with significant accuracy, from simple measures (questionnaire-based interoceptive awareness and sleep quality).

Turning to the weaknesses of our study, our recruitment procedure was unconstrained, i.e., we did not pre-select PwMS on the basis of any criteria. On the one hand, this is a strength as we avoided recruiting a specific subgroup that could have led to a biased perspective. On the other hand, there are probably multiple pathophysiological mechanisms that lead to fatigue in MS (Manjaly et al., 2019), and, given this likely heterogeneity and the absence of data on interoception and metacognition in MS prior to the start of our study, it was not possible to determine an adequate sample size. Second, novel methods for assessing metacognition of interoception have been introduced only very recently (Harrison, Garfinkel, *et al*., 2021; Nikolova *et al*., 2022; Legrand *et al*., 2022) and were not available when our study started. However, we acknowledge that such a task-based measure of interoceptive accuracy could provide relevant data regarding the correspondence between objective interoception and participants’ beliefs about their reported interoception (instead, we relied on a well-validated questionnaire of interoceptive awareness). It is worth mentioning that even these task-based procedures do not yet allow for assessing the particular metacognitive construct of interoception that the ASE theory focuses on, i.e. self-monitoring of the brain’s capacity to control bodily states. We therefore had to resort to an indirect approach, using a task that probes metacognition about exteroception (specifically, confidence about perceptual decisions in the visual domain). This was motivated by a recent study (Rouault et al., 2018) showing that metacognitive bias (confidence level) during this specific task was associated with apathy. However, apathy is not a fully identical construct and shares both similarities and differences with fatigue (Daumas *et al*., 2022). Indeed, in our sample, we could not detect an association between metacognitive bias and fatigue. One potential reason could be statistical power: the sample of PwMS in this study was much smaller than the general population sample from Rouault et al. (2018). Additionally, according to the ASE theory, a link between fatigue and exteroceptive metacognition is only to be expected once a generalisation of low self-efficacy beliefs has taken place – a process that, according to the theory, should be reflected by the onset of depression.

This directly leads us to the third, and most important, limitation of the current study: our particular sample of PwMS did not exhibit a particularly high degree of depressive symptoms, which is not congruent with a key assumption inherent to the ASE theory. This observation represents an important caveat for all analyses of exteroceptive metacognition presented in this study. More specifically, according to the ASE theory, alterations of exteroceptive metacognition are only expected to occur once a generalisation of low self-efficacy beliefs, manifesting as depression, has taken place (Stephan et al. 2016). The fact that only a small subgroup of our participants was found to exhibit notable depressive symptoms casts doubt on whether our particular sample is well suited to test for significant metacognitive changes in the exteroceptive domain. This doubt is strengthened further by the observation that, in our sample, there is no association between local (task-based) confidence level and global confidence (self-efficacy). By contrast, the questionnaire-based and physiological assessments are not affected by this potential problem since they provide measures unrelated to the exteroceptive domain and do not rely on the assumption that generalisation of low self-efficacy beliefs having taken place.

Notwithstanding these weaknesses, our study makes several important contributions to a better understanding of fatigue in MS. In particular, our results support the notion that interoception is an important factor for fatigue and demonstrate the feasibility of predicting individual levels of fatigue from simple questionnaire-based measures not directly related to fatigue. In future work, we will aim to replicate these findings in larger samples and address the important challenge of developing experimental procedures that allow for assessing metacognition of interoceptive processes. We hope that this work will eventually lead to clinically useful procedures of differential diagnosis that help identifying patients who would benefit from cognitive interventions targeting interoception and metacognition (Manjaly et al. 2019).

## Acknowledgments

We would like to thank Jakob Heinzle for helpful discussions about statistical analyses, Fabien Vinckier for early discussions about the protocol design, Nathalie Appenzeller for help with data checks, and the Swiss Multiple Sclerosis Register for assistance with recruitment.

MR is the beneficiary of a postdoctoral fellowship from the AXA Research Fund. KES acknowledges funding by the René and Susanne Braginsky Foundation and the University of Zurich. ZMM acknowledges support by the Wilhelm Schulthess Foundation, the Olga Mayenfisch Foundation, and the Swiss MS Society.

MR work was also supported by La Fondation des Treilles, and by a department-wide grant from the Agence Nationale de la Recherche (ANR-17-EURE-0017, EUR FrontCog). This work has received support under the program «Investissements d’Avenir» launched by the French Government and implemented by ANR (ANR-10-IDEX-0001-02 PSL).

## Author contributions

Conceptualization: MR, SF, KES, ZMM

Data Collection: ZMM, IP

Methodology: MR, HG, SF, KES

Formal Analysis: MR, HG

Writing – Original Draft: MR, KES, ZMM

Writing – Review & Editing: MR, SF, IP, HG, KES, ZMM

Supervision: KES, ZMM

Funding Acquisition: KES, ZMM

## List of abbreviations

ASE: allostatic self-efficacy
DDM: drift-diffusion model
FSS: fatigue severity scale
GLM: general linear model
GSES: general self-efficacy scale
HADS: hospital anxiety and depression scale
HRV: heart rate variability
MAIA: multidimensional assessment of interoceptive awareness
MFIS: modified fatigue impact scale
MS: multiple sclerosis
MSSE: multiple sclerosis self-efficacy scale
PCA: principal component analysis
PEs: prediction errors
PwMS: persons with multiple sclerosis
PSQI: Pittsburgh sleep quality index
SD: standard deviation
SSR: sympathetic skin response

