## Supplementary Information for "Interoceptive and metacognitive facets of fatigue in multiple sclerosis"

**Analysis A: Is fatigue related to measures of interoception and autonomic regulation?**

In this section, we report several analyses that go beyond our preregistered analysis plan and examine potential reasons why Analysis A failed to find the expected association between fatigue levels and measures of autonomic function. In particular, we examined the possibility that using the first principal component as a summary of the various autonomic measurements may have been an inadequate choice.

Given that the structure of the principal components did not show the rapid decay of eigenvalues we had expected and PC1 therefore accounted for only a relatively moderate amount of variance (Fig. 4B), we reasoned that adding the second principal component (PC2) to the regression model might result in a more appropriate representation of the variance structure of physiological measurements. Therefore, we repeated the analysis using both PC1 and PC2 as regressors. Again, we found that self-report measures of interoception ($t_{52}$=-2.75, $p$=0.0087 uncorrected, $p$=0.016 FDR-corrected) significantly explained fatigue (MFIS) scores, whereas none of the two principal components showed a significant relationship to fatigue (PC1: $t_{52}$=0.14, $p$=0.89; PC2: $t_{52}$=1.44, $p$=0.16). For completeness, instead of relying on PC1 (or PC1 and PC2) from our PCA analysis (Fig. 4), we repeated this analysis using each physiological regressor separately. In all cases, consistent with the above results, we found no significant contribution of any of the six physiological regressors when entered in isolation (all $t_{52}$<1.45, all $p$>0.15) to explaining fatigue (MFIS) scores. For completeness, to assess whether fatigue scores were explained by any linear combination of physiological regressors, we also examined additional models in which all of the six physiological regressors are entered together. We found that the model explained significant variance in FSS (F-test: $p$=0.0033; N=63 participants), but not MFIS (F-test: $p$=0.0786; N=53 participants) scores. However, none of the six physiological regressors contributed significantly to explaining MFIS scores (all abs($t_{52}$)<1.59, all $p$>0.12), while again the MAIA (combined subscales 3 and 8) did ($t_{52}$=-2.92, one-tailed $p$=0.0028). Likewise, none of the six physiological regressors contributed significantly to explaining FSS scores (all abs($t_{52}$)<1.63, all $p$>0.11), while again MAIA ($t_{52}$=-2.98, $p$=0.0045) did.

The above control analyses examined the association of individual autonomic measurements to fatigue scores. In three additional analyses, we examined whether fatigue scores could be explained by a linear combination of physiological regressors. To this end, we first revisited the GLM containing all physiological regressors (in addition to MAIA and confirmed regressors), conducting an F-test for the joint effect of all six physiological regressors. This provided a nonsignificant result (F-test from the overall model but restricted to the physiological regressors: $p$=0.2297 for MFIS, $p$=0.5003 for FSS). Second, we applied a GLM containing only the six physiological regressors (without MAIA or confound regressors of no interest). We reasoned that correlations between MAIA scores and the physiological regressors (Fig. S1) might have masked links between physiological regressors and fatigue in the above analyses. However, none of these models explained a significant amount of variance (F-tests: $p$=0.3869 for MFIS, $p$=0.1235 for FSS). Third, for the sake of completeness, we constructed a GLM with the six physiological regressors and confound regressors, only leaving out the MAIA regressor. For MFIS, an F-test for the whole model was nonsignificant ($p$=0.4339), nor was an F-test for the six physiological regressors only ($p$=0.7027). For FSS, an F-test for the whole model was significant ($p$=0.0341), but the F-test for the six physiological regressors was not ($p$=0.9332).

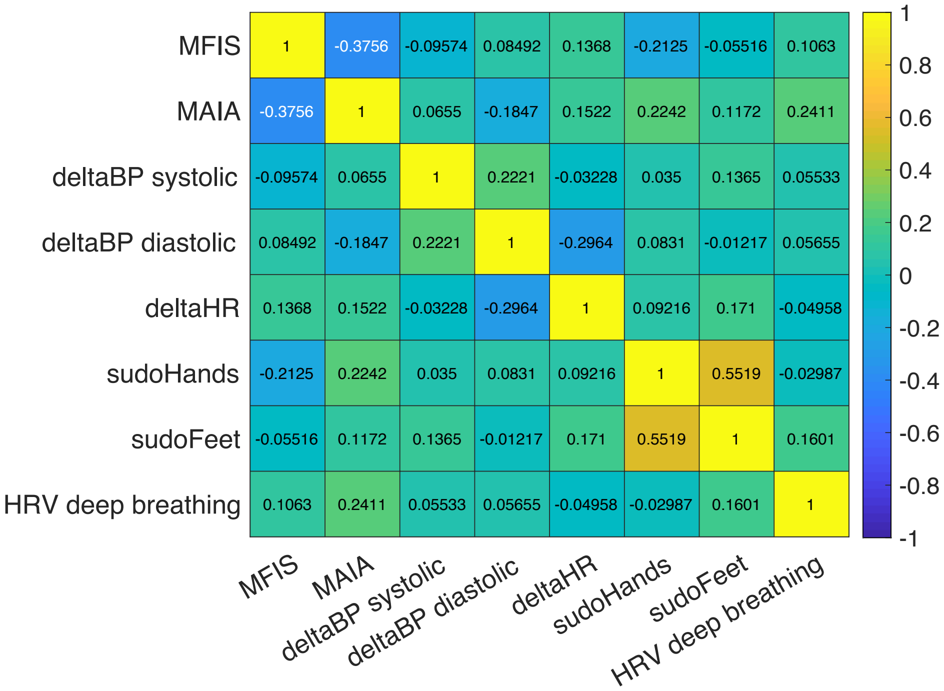


***Figure S1:*** *Matrix of pairwise correlations between fatigue scores (MFIS), MAIA (combined subscales 3 and 8), and physiological measurements: heart rate variability (HRV, computed as RMSSD during deep breathing), systolic and diastolic ΔBP and ΔHR (standing up after resting in supine position for 10 minutes), and sudomotor activity (hands and feet).*

### Elastic net regression

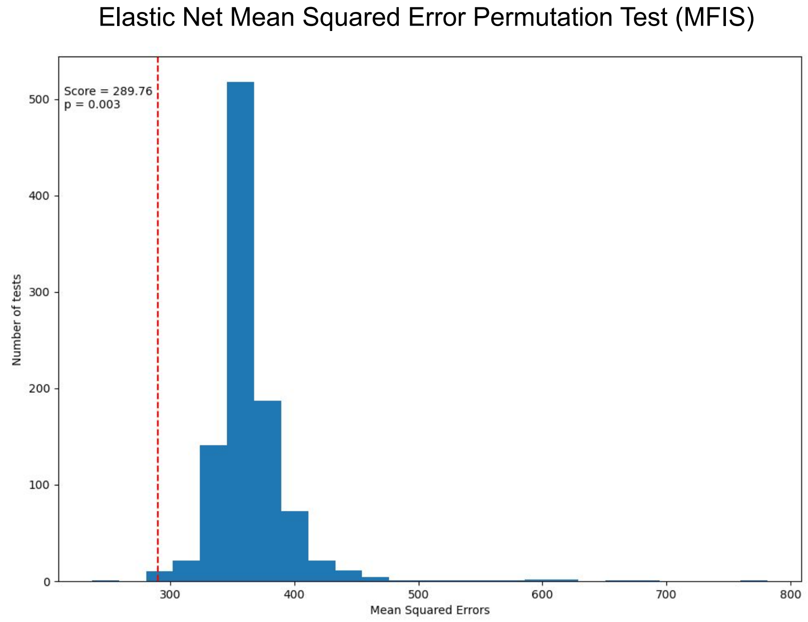


***Figure S2:*** *Null distribution (based on mean squared error, MSE) for the elastic net regression model predicting MFIS fatigue scores (N=52) from all 15 measurements. Dashed red line shows the MSE of the unpermuted prediction. Blue bars show the MSE distribution (N=999) when permuting scores.*

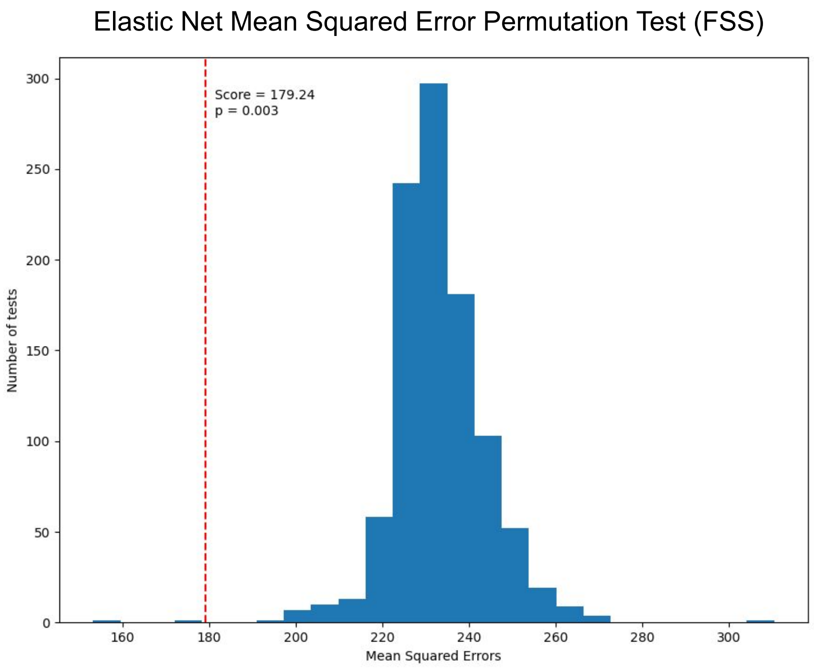


***Figure S3:*** *Null distribution (based on mean squared error, MSE) for the elastic net regression model predicting FSS fatigue scores (N=62) from all 15 measurements. Dashed red line shows the MSE of the unpermuted prediction. Blue bars show the MSE distribution (N=999) when permuting scores.*
